## Supplementary Appendix for "Impact of Covid-19 social distancing measures on future incidence of invasive pneumococcal disease in England and Wales – a mathematical modelling study"

Table S1. Cumulative difference in cases of invasive pneumococcal disease over 5 epidemiological years (July to June) starting from 2019/2020 by age group with different levels of reduction in PCV13 coverage and in contact rates during two lockdown periods starting 23 March for two months and 5 November 2020 for six months. Results show median and minimum and maximum of the range of model outputs generated by 500 model parameter sets [1]

| Contact rate reduction | Vaccine coverage reduction | <2 yrs | 2-4 yrs | 5-14 yrs | 15-44 yrs | 45-64 yrs | 65+ yrs | Total |
|---|---|---|---|---|---|---|---|---|
| 0% | 40% | 3 (2, 5) | 0 (0, 1) | 0 (0, 1) | 4 (2, 7) | 4 (2, 7) | 6 (2, 13) | 18 (8, 33) |
| | 100% | 12 (6, 20) | 2 (1, 3) | 1 (1, 2) | 16 (9, 26) | 17 (8, 29) | 25 (9, 50) | 73 (33, 130) |
| 10% | 0% | -130 (-161, -97) | -47 (-57, -33) | -79 (-102, -74) | -617 (-697, -577) | -1021 (-1243, -890) | -2136 (-2484, -1837) | -4032 (-4489, -3606) |
| | 40% | -128 (-159, -95) | -47 (-57, -33) | -79 (-102, -74) | -615 (-693, -575) | -1018 (-1240, -887) | -2130 (-2480, -1834) | -4027 (-4482, -3604) |
| | 100% | -122 (-154, -88) | -46 (-56, -32) | -78 (-101, -73) | -607 (-685, -568) | -1009 (-1230, -880) | -2118 (-2459, -1825) | -3991 (-4432, -3566) |
| 40% | 0% | -471 (-580, -348) | -169 (-208, -118) | -275 (-349, -256) | -2062 (-2292, -1915) | -3368 (-4062, -2931) | -7107 (-8135, -6201) | -13494 (-14686, -12221) |
| | 40% | -471 (-578, -347) | -169 (-208, -118) | -275 (-349, -256) | -2061 (-2291, -1913) | -3367 (-4062, -2931) | -7107 (-8135, -6201) | -13494 (-14676, -12211) |
| | 100% | -468 (-576, -345) | -169 (-207, -117) | -275 (-349, -256) | -2059 (-2288, -1912) | -3366 (-4062, -2930) | -7107 (-8125, -6201) | -13484 (-14676, -12211) |
| 74% | 0% | -618 (-768, -447) | -221 (-280, -152) | -356 (-453, -329) | -2626 (-2890, -2393) | -4279 (-5183, -3690) | -9033 (-10248, -7910) | -17209 (-18511, -15541) |
| | 40% | -617 (-768, -446) | -221 (-280, -152) | -356 (-453, -329) | -2626 (-2890, -2393) | -4279 (-5183, -3690) | -9033 (-10248, -7910) | -17209 (-18511, -15540) |
| | 100% | -617 (-767, -446) | -221 (-280, -152) | -356 (-453, -329) | -2626 (-2890, -2392) | -4279 (-5183, -3690) | -9033 (-10248, -7910) | -17189 (-18511, -15540) |

Table S2. Simulation results for the number of overall IPD cases between February and June 2020 and proportional reduction compared to the model predictions for the same period in 2019 (2,350 (UI 2,129, 2,531) IPD cases) in England and Wales according to different reduction levels in contact rates and vaccine coverage during two lockdowns due to COVID-19 in the UK. Results show median and minimum and maximum of the range of model outputs generated by 500 model parameter sets [1]

| Contact rate reduction | Vaccine coverage reduction | IPD cases February to June 2020 | Proportional reduction compared with February to June 2019 |
|---|---|---|---|
| 0% | 40% | 2,374(2,146, 2,554) | -0.9%(-1.3%, -0.6%) |
| | 100% | 2,375(2,148, 2,555) | -1.0%(-1.3%, -0.7%) |
| 10% | 0% | 1,953(1,766, 2,101) | 16.9%(16.5%, 17.3%) |
| | 40% | 1,954(1,766, 2,102) | 16.9%(16.5%, 17.3%) |
| | 100% | 1,955(1,767, 2,103) | 16.9%(16.5%, 17.2%) |
| 40% | 0% | 1,125(1,017, 1,211) | 52.2%(51.8%, 52.4%) |
| | 40% | 1,125(1,017, 1,211) | 52.1%(51.8%, 52.4%) |
| | 100% | 1,126(1,018, 1,212) | 52.1%(51.8%, 52.4%) |
| 74% | 0% | 677(612, 728) | 71.2%(71.1%, 71.3%) |
| | 40% | 677(612, 728) | 71.2%(71.1%, 71.3%) |
| | 100% | 677(612, 729) | 71.2%(71.1%, 71.3%) |

**Model description**

Choi et al. [1] developed an age-structured, deterministic model structure to describe the pneumococcal transmission dynamics in England and Wales in order to estimate the potential impact of revising the 13-valent pneumococcal conjugate vaccine schedule from 2+1 to 1+1 in England and Wales. The data sources, model description, parameter estimation and fitting procedures described in this supplementary Appendix are a summary of those from Choi et al. [1]

***Serotype groupings***

In the model serotypes are grouped according to whether they are covered by the seven-valent pneumococcal conjugate vaccine (PCV7, VT1), or are one of the additional serotypes covered by PCV13 (VT2) or are non-vaccine serotypes (NVT). As in Choi et al [1] serotype 1 was excluded from the VT2 group as its incidence fell progressively after introduction of PCV7 in 2006 even though serotype 1 was not covered by PCV7, indicating secular changes in the behaviour of this serotype that are unrelated to vaccination. Further reductions in serotype 1 occurred after PCV13 introduction with continuing very low incidence [1] ; its exclusion from the model will therefore have a negligible impact on future IPD incidence. Serotype 3, which is also covered by the additional serotypes in PCV13, has increased progressively since 2013/14 behaving like a non-PCV13 serotype that exhibits serotype replacement. It was therefore included in the model in the NVT group [1]. For the current analysis, vaccine serotype outputs are shown combined as a single VT group though in the fitting stage VT1 and VT2 are separated to reflect the different invasiveness potential and propensity to induce serotype replacement of the VT1 and VT2 serotypes, and the differences between PCV7 and PCV13 vaccines in protecting respectively against carriage acquisition of VT1 and VT2 serotypes.

***Model data***

The datasets used for this modelling study include carriage prevalence data during the pre-PCV era, annual population changes by age, annual IPD cases by age groups and serogroups in pre- and post PCV introduction up to 2015/16 in England and Wales, PCV coverage data, and contact patterns in the population.

#### 1. *Carriage prevalence and IPD incidence*

The pre-PCV7 carriage prevalence in seven age groups (under 1, 1-2, 3-4, 5-9, 10-19, 20-39 and 40+ years) and the three serotype groupings VT1, VT2 and NVT was obtained from a longitudinal family study [2] in which 3,869 nasopharyngeal swabs were taken from 489 individuals who were swabbed monthly over ten months between 2001 and 2002 in England.

The national enhanced surveillance dataset of serotyped IPD cases collated by Public Health England for epidemiological years (July to June) between 2000/01 and 2005/6 was used for the pre-PCV7 baseline incidence and the epidemiological years from 2005/6 to 2015/16 as the post-PCV period for the purposes of model fitting. The carriage prevalence and IPD datasets are available at https://doi.org/10.1371/journal.pmed.1002845.s001.

#### 2. *Population structure and contact patterns*

The annual population size by age in each year from the pre-PCV7 to the post-PCV13 period was obtained from census data and the estimated demographic changes out to 2030 from the Office of National Statistics [3]. The contact pattern within and between age groups was derived from the POLYMOD survey conducted in the UK in 2006 [4] supplemented by an additional contact survey among infants under one year [5].

#### 3. *PCV coverage data*

Vaccine coverage by dose, monthly birth cohort and calendar month in the PCV7 catch-up and the routine 2+1 PCV7 programme up to 2008 was obtained from the General Practice Research database as previously described [6]. Thereafter, coverage for the second priming dose and the booster dose was obtained from annual national coverage data [7].

***Model structure***

#### 1. *Model population*

The population in the model is divided into 100 annual age cohorts (0, 1, 2, 3, … , 99). Each annual age-cohort is divided into 48 equal-sized age-cohorts (in total 4,800 age-cohorts in the total population in the model).

#### 2. *Transition between model compartments*

In the absence of vaccination individuals are born susceptible (S) to pneumococcal carriage and become infected (I) with a VT1, VT2 or NVT serotype as determined by the serogroup-specific force of infection. An episode of carriage does not result in protection against subsequent carriage of any serotype (i.e. SIS model structure). Invasive disease is assumed to occur at the time of carriage acquisition whereas transmission can occur at any time during the carriage episode. Individuals clear their infection with age-dependent clearance rates as estimated previously [8] and become susceptible again. Individuals already carrying a serotype from one of the three groupings will have some degree of protection against infections from another serotype grouping according to the level of competition between the three serotype groupings. These competition parameters determine the extent of serotype replacement in carriage (and therefore in IPD) when vaccine serotypes decline post-PCV introduction. The flowchart for the model structure is available at https://doi.org/10.1371/journal.pmed.1002845.g002.

#### 3. *Vaccine efficacy parameters*

Within the vaccine protected group, two doses in the first year of life or one dose after 12 months of age are assumed to confer the maximum degree of protection that can be obtained from PCV7 or

PCV13  against carriage acquisition of the respective serotype grouping (termed full protection). A single dose in the first year of life is assumed to provide half the maximum protection (termed partial protection). Protection wanes exponentially with fully protected individuals moving back to the partially protected group and then back to the fully susceptible group. For the base case the average duration of protection for both full and partial protection is set at 5 years.

Vaccine efficacy against IPD given carriage of a vaccine serotype for fully protected individuals is assumed to be 100% at the time of vaccination and to wane with the same average duration as for the full and partial protection respectively against carriage.  The flowchart showing the transitions between the unvaccinated, partially protected and fully protected compartments as a result of vaccination and subsequent waning is available at https://doi.org/10.1371/journal.pmed.1002845.g003.

The propensity to develop invasive disease upon carriage acquisition within each age group will be determined by the overall case-carrier ratio (CCR) of serotypes comprising the VT1, VT2 and NVT groupings (https://doi.org/10.1371/journal.pmed.1002845.g006) and is derived from the ratio of IPD cases: incident carriage infections within each serotype grouping in each age group.

***Model parameter estimation***

The model fitting process consists of a static pre-vaccination equilibrium component and post-vaccination dynamic component. By fitting the model to the given datasets, we estimate 26 model parameters: competition parameters for six age groups and three serotype groupings,  the two vaccine efficacies against carriage acquisition of VT1 and VT2  serotypes  and an additional parameter that allows for an increase in the case carrier ratio of the serotypes in the NVT group after 2013/14 when the rate of serotype replacement suddenly increased [9]. This last parameter is allowed to vary between each of the six age groups generating in total 26 parameters.  A summary table showing the derivations of the model parameters is available at https://journals.plos.org/plosmedicine/article/figure?id=10.1371/journal.pmed.1002845.t001.

As described in Choi et al [1] the force of infection and clearance parameters that determine the rate of acquisition and termination of a new carriage episode for the VT1, VT2 and NVT groups were estimated by fitting a static model to the pre-vaccination carriage data. The competition parameters that determine the extent to which carriage of serotypes in one group protects against acquisition of serotypes in another group were estimated by fitting a post-vaccination model to the changes in serotype grouping after the introduction of PCV7 and PCV13 from 2005/6 to 2015/16.

The uncertainty intervals were generated from 500 randomly selected sets of model parameters that generated outputs within ±0.3 of the set that gave the maximum likelihood value obtained using the Nelder-Mead Downhill Simplex method [10] when compared with the post-PCV IPD data. Results are shown as the median value of the accepted 500 parameters sets and the minimum to maximum range is shown as the uncertainty interval (UI) at https://journals.plos.org/plosmedicine/article/figure?id=10.1371/journal.pmed.1002845.t002. VEcVT1 (Vaccine efficacy against acquiring VT1 carriage) and VEcVT2 (Vaccine efficacy against acquiring VT2 carriage) were estimated at 0.55 (0.53, 0.57) and 0.30 (0.26, 0.36) respectively (minimum and maximum).

***Long-term simulations***

Using the 500 parameter sets obtained for the uncertainty boundary the model simulated various scenarios to investigate the potential impact of two lockdowns occurred during the COVID-19 pandemic until 2030/31, the furthest year for which there are population age-structure predictions.

### References

1 Choi YH, Andrews N, Miller E. Estimated impact of revising the 13-valent pneumococcal conjugate vaccine schedule from 2+1 to 1+1 in England and Wales: A modelling study. *PLoS*

*Med* 2019;**16**. doi:10.1371/journal.pmed.1002845

2 Hussain M, Melegaro A, Pebody RG, *et al.* A longitudinal household study of Streptococcus pneumoniae nasopharyngeal carriage in a UK setting. *Epidemiol Infect* 2005;**133**:891–8.

3 Office for National Statistics. England Wales population size. https://www.ons.gov.uk/

4 Mossong JJ, Hens N, Jit M, *et al.* Social contacts and mixing patterns relevant to the spread of infectious diseases. *PLoS Med* 2008;**5**:e74.

5 van Hoek AJ, Andrews N, Campbell H, *et al.* The social life of infants in the context of infectious disease transmission; social contacts and mixing patterns of the very young. *PLoS One* 2013;**8**:e76180.http://www.pubmedcentral.nih.gov/articlerender.fcgi?artid=3797797&tool=pmcentrez&rendertype=abstract (accessed 30 Sep 2014).

6 Choi YH, Jit M, Gay N, *et al.* 7-Valent Pneumococcal Conjugate Vaccination in England and Wales: Is It Still Beneficial Despite High Levels of Serotype Replacement? *PLoS One* 2011;**6**:e26190.

7 Public Health England. Vaccine uptake guidance and the latest coverage data. 2018.https://www.gov.uk/government/collections/vaccine-uptake

8 Melegaro A, Gay NJ, Medley GF, *et al.* Estimating the transmission parameters of pneumococcal carriage in households. *Epidemiol Infect* 2004;**132**:433–41.

9 Ladhani SN, Collins S, Djennad A, *et al.* Rapid increase in non-vaccine serotypes causing invasive pneumococcal disease in England and Wales, 2000-17: a prospective national observational cohort study. *Lancet Infect Dis* 2018;**18**:441–51.

10 Nelder JA, Mead R. A Simplex Method for Function Minimization. *Comput J* 1965;**7**:308–13. doi:10.1093/comjnl/7.4.308
